# Differences in Cardiovascular Disease Burden, Screening, Education, and Care by Clinic Type in the 2022 Health Center Patient Survey

**DOI:** 10.64898/2026.04.14.26350912

**Authors:** B King, B Beech, O Jones, E Castillo, S Attri, D Buck

**Affiliations:** University of Houston, Tilman J Fertitta Family College of Medicine, Department of Health Systems and Population Health Sciences; Houston, TX, USA; University of Houston, Humana Integrated Health Systems Sciences Institute; Houston, TX, USA; University of Houston, Office of Population Health; Houston, TX, USA; University of Texas Health Science Center at Houston, School of Public Health; Houston, TX, USA; University of Texas at Austin, Public Health Program; Houston, TX, USA; University of Houston, Tilman J Fertitta Family College of Medicine, Office of Community Health; Houston, TX, USA

## Abstract

**Background:** Persons experiencing homelessness (PEH) have a 2-3-fold greater risk for cardiovascular disease (CVD) mortality compared with domiciled counterparts. Evidence has repeatedly shown elevated chronic disease burden, reduced access to many types of care, and lower utilization of medication to control CVD risk factors in clinical settings dedicated to providing health care to PEH. There are federally funded health clinics targeting barriers to access for patient populations experiencing homelessness in place. These clinics are frequently overwhelmed and limited by their scope to primary care despite well documented burdens of co- and tri-morbid conditions. There is scarce evidence on differences between access, quality, and experiences of care delivered relative to other safety-net models.

**Method:** The 2022 Health Center Patient Survey (HCPS) was collected on behalf of the Health Resources and Services Administration (HRSA). The HCPS is a nationally representative, three-staged, sample-based survey collected via 1:1 interview with clinic patients. The survey assessed sociodemographics, health conditions and behaviors, access to and utilization of care, and patients’ experiences with comprehensive services they received at HRSA-funded Federally Qualified Health Centers (FQHCs), including community health centers (CHC), healthcare for the homeless (HCH) clinics, and public housing primary care (PHPC) clinics. One hundred and three unique awardees and 318 health center sites were recruited, and 4,414 patient interviews were completed. Investigators analyzed patient characteristics and multiple survey items related to AHA’s *Essential 8* metrics for differences between HCH and CHC patient responses.

**Results:** HCH clinics had fewer elderly patients (∼7%) than CHCs (∼17%). Reported 7-day physical activity measures, average sleep below 7 hours per day, and Lifetime smoking (>100 cigarettes; OR=4.2, p<0.001) were all greatest among HCH patients. Fewer HCH patients reported ever having or recent lipid tests (both p<0.001). HCH patients were more likely to report hypertension (p=0.003) but less likely to report receiving nutrition advice (all p<0.05). HCH patients were less likely to be taking medication even if it was prescribed (p<0.001). Adjustments for differences in age or CVD history were able to explain some observed differences but increased the magnitude of other disparities.

**Conclusions:** CVD burden differs across the various HRSA funding mechanisms for clinics, as do demographics and multiple metrics of health behaviors and biomarkers of cardiovascular health. Greater disease burden in HCH patients is likely compounded by increased risk factors and underperformance in providing health education interventions.

**Clinical Perspective:** *What Is New?:*

- Patients accessing Health Care for the Homeless clinics demonstrate unique cardiovascular risk profiles characterized by higher rates of inadequate sleep, smoking history, and pre-diabetes compared to Community Health Center patients, even after adjusting for sociodemographic factors.

*What Are the Clinical Implications?:*

- Traditional cardiovascular disease risk assessment tools and prevention strategies may need to be recalibrated for homeless populations, as standard clinical metrics and screening approaches may not fully capture the complex interplay of behavioral, environmental, and social exposures affecting this vulnerable group.

*Research Perspective:* What New Question Does This Study Raise?

- How do structural inequities and comorbid conditions resulting in and from homelessness impact health in ways that may not be captured by conventional risk assessment tools? What Question Should be Addressed Next?

- What modifications to evidence-based cardiovascular interventions are needed to effectively serve people experiencing homelessness, and how can these interventions be integrated into Health Care for the Homeless clinics and other FQHCs?

## Introduction

Cardiovascular Disease (CVD) is the leading cause of global mortality and is a major contributor to age-adjusted disability ^1^. In the United States, it is also the leading cause of death for people of most racial/ethnic groups, with an estimated cost of >$200 billion annually in healthcare services, medications, and lost productivity ^2^. In 2022, the American Heart Association (AHA) published *Life’s Essential 8* (LE8) for defining ideal Cardiovascular Health (CVH) ^3^. These eight domains include 1) Diet, 2) Exercise, 3) Smoking Cessation, 4) Sleep, 5) Weight Management and management of 6) Cholesterol, 7) Blood Sugar, and 8) Blood Pressure ^4^ (BP). Numerous studies have shown that improving these CVH domains leads to a lower risk of Cardiovascular Disease (CVD) ^5–8^.

Non-pharmacological management of these diseases are also encompassed in the LE8. Endeavors toward tobacco cessation, weight loss, diet (DASH diet, sodium restriction, increased vegetable intake, minimally processed carbohydrates, etc.), and exercise have all been shown to improve these chronic diseases. They are often indicated either as first-line management or in conjunction with pharmacological management, which can be implemented to help achieve better control of these diseases ^2,9–11^.

Federally Qualified Health Centers (FQHCs) are community-based healthcare organizations providing primary care services to underserved populations. These organizations qualify for increased federal reimbursements from the Health Resources and Services Administration (HRSA) to serve disadvantaged populations ^12^. Many uninsured and medically underserved individuals seek care through FQHCs ^13–19^, including general Community Health Centers (CHCs), and targeted clinic funding for health care for the homeless (HCH) clinics.

These HCH facilities are particularly important to providing healthcare to people experiencing homelessness (PEH). PEH have a lifespan that is 17.5 years shorter than that of the general population ^20^. The most frequent causes of death in these individuals are deaths from circulatory system diseases (33.8%) ^20^, which is also the case for the general population with cardiovascular disease. These deaths are likely due, in part, to increased risk factors from AHA’s LE8, including increased rates of smoking ^21^, lack of a safe home and therefore lack of sleep ^22^, poor diet ^23^, and subpar management of chronic diseases related to CVD risk; hyperlipidemia ^24^, diabetes ^25^, and hypertension ^26^.

PEH must prioritize basic needs, such as finding shelter and food, over getting health and social care. Bureaucratic structures, rigid opening hours, and discrimination and stigma, hinder these individuals’ access to health and social care ^27^. PEH had worse health and were twice as likely to have unmet medical needs. They were also more likely to need prescriptions and have unmet needs related to receiving prescriptions. They were also twice as likely to have an ED visit in the past year. Homeless patients more often reported that their health center provided enabling services, including free medication ^28^. PEH tend to have worse health; 76% have at least one unmet healthcare need; 32% are unable to obtain needed medical or surgical care, 36% need prescription meds, 21% have mental health issues, 41% need eyeglasses, and 41% require dental care ^29^. Homeless people have higher rates of premature mortality than the rest of the population, especially from suicide and unintentional injuries, and an increased prevalence of a range of infectious diseases, mental disorders, and substance misuse ^30^.

If PEH experience successful primary care that may result in lifesaving measures with lower costs to hospital systems ^31^. Overall, individuals seeking care at any type of FQHC tend to be more disadvantaged and in poorer health; however, they may be more likely to use preventative care than those without access ^14^. Patients in CHCs reported high-quality primary care experiences, especially regarding accessibility and communication ^19^. Compared to non-HRSA-funded safety net clinics, FQHC patients had lower likelihood of unmet medical and dental needs, potentially because FQHCs are seen as accessible, affordable, and able to provide comprehensive care ^32^.

Although these facilities are hallmark locations for care for underserved individuals, seeking care through an FQHC may also be associated with delays in access to medication and continuity of disease management ^33^. Approximately 29% of participants with type 2 diabetes had delayed access, and 24% were unable to get medications ^33^. Barriers to care included personal barriers, such as federal poverty level, health status, and psychological distress, all associated with being unable to get medications. Financial barriers, including out-of-pocket medication costs and employment, were associated with access to prescription medications. Critically, the type of health center funding program as a structural barrier was also associated with access to medications. Receiving care from HCH or Public Housing Primary Care (PHPC) clinics was associated with delayed medication access ^33^. Many patients seeking care at FQHCs for asthma treatment also reported experiencing delays, resulting in more frequent asthma exacerbations and hospital/emergency visits due to asthma-related issues ^34^. Despite FQHCs providing care for this population to bridge this primary care gap, other evidence has shown that seeking care from a homeless shelter specifically (alone or with an FQHC) may also result in lower medication adherence ^35^.

Furthermore, there is conflicting evidence on the efficacy of chronic disease management for patients at FQHCs. One study found lower rates of cholesterol treatment discussion and statin prescriptions for CHC clinic patients, but that that single risk factor management approaches strongly influenced both process metrics ^24^. Another study found slightly lower rates of most diabetes care process measures, higher rates of ED visits, but significantly decreased likelihood of hospitalization compared to non-FQHC patients ^25^. On the other hand, hypertension management in Medicaid-covered individuals at CHCs was found to be similar to that of private physician offices, but with less usage of fixed-dose combination drugs at CHCs despite higher rates of uncontrolled hypertension at initial presentation ^26^. For FQHCs, adopting a patient-centered medical home approach to disease management results in better hypertension coaching and management and reduced ED visits ^13^. The inclusion of pharmacists increased patient relationships and trust, as well as medication adherence ^36^. Furthermore, pharmacist inclusion also resulted in better control of diabetes and hypertension ^37^. Introduction of a Hypertension Management Program, which included a multidisciplinary team approach to patient hypertension management, resulted in increases in hypertension control ^38^.

Lack of insurance remains a potential obstacle to accessing healthcare, even in FQHCs, where a patient’s ability to pay is not an allowable exclusion for care ^16^. Some national studies have demonstrated that disadvantaged individuals still feel that lack of health insurance remains a large barrier to care ^29^.Compared to those covered by Medicaid, uninsured individuals had fewer physician visits, demonstrating unmet need, and were less likely to be able to fill prescriptions ^39^. Patients without insurance had less favorable experiences than their insured counterparts while seeking care within CHC, especially regarding the comprehensiveness and coordination ^19^. Uninsured patients were more likely to have barriers/delays than private insurance and Medicare/Medicaid patients ^40^. Additionally, those who are uninsured usually have poorer chronic disease management, especially regarding HTN and HLD ^41^.What’s more, evidence has shown that 5 years after Medicaid expansion, there were improvements in BP and glucose control. Patients who gain healthcare coverage were more likely to use primary care services provided by FQHCs ^16^. And yet, cardiovascular risk factor modification is suboptimal among homeless adults in Toronto despite universal health insurance. A study in that international setting showed that multiple risk factor equations may underestimate true risk in this population because of inadequate accounting for factors such as cocaine use and heavy smoking ^5^.

There is also a paucity of evidence for any differences in delays between different social groups seeking care at FQHCs. Some data suggest no significant difference in care between racial/ethnic groups ^19^.However, other data suggest that being Hispanic or black was protective of delays/barriers to care ^40^. Minorities were more likely to receive diet counseling, reduce their sodium intake, and adhere to exercise counseling. Some were less likely to take hypertension control medication and more likely to have hypertension-related ED visits ^42^. There were fewer hospitalizations for ambulatory care-sensitive conditions among blacks and Hispanics who used these health centers, and use of health centers was also associated with 3% and 12% fewer hospitalizations for ambulatory care-sensitive conditions among nonelderly disabled blacks and Hispanics, respectively ^15^.

The purpose of this investigation is to compare patient characteristics and multiple survey items related to AHA’s *Essential 8* metrics for differences across the CHC and HCH clinic patients. As mentioned, cardiovascular disease prevention involves a multifaceted approach to both social and medical risk factors. Because of the limited access to clinics and medications, an improved understanding of the differences in care management between these outpatient populations is key. This study highlights the differences in chronic disease management of HTN, HLD, and DMT2 to evaluate if improvements can be made in this vulnerable patient population.

## Method

This study reflects a secondary analysis of the public-use file of the 2022 Health Center Patient Survey (HCPS) collected by the Health Resources and Services Administration (HRSA). This was a cross-sectional survey of patients seen in two types of HRSA-funded clinics: CHCs and HCH clinics. The HRSA Health Center Program primarily serves medically underserved populations who often face significant barriers to healthcare access, including individuals with low income, racial and ethnic minorities, uninsured or underinsured patients, agricultural workers, residents of public housing, and people experiencing homelessness. These clinics provide comprehensive primary care services, often functioning as a critical access point for vulnerable groups who might otherwise remain disconnected from the healthcare system.

### Sample

The 2022 HCPS employed a rigorous three-stage sampling design to produce a nationally representative sample of patients served by the Health Center Program. In the first stage, health center grantees were selected using stratified sampling based on key characteristics, including patient demographics (e.g., race/ethnicity), grantee size, and funding type (e.g., Health Care for the Homeless or Public Housing Primary Care programs). In the second stage, three or more delivery sites were randomly selected within each sampled grantee. In the final stage, a random sample of patients was drawn at each participating site, resulting in in-person interviews conducted with a representative subset of patients aged 18 and older.

To ensure adequate representation of historically underserved groups, the HCPS employed oversampling of specific racial and ethnic populations, including Asian, American Indian/Alaska Native, and Native Hawaiian/Pacific Islander patients. This strategy was designed to support more granular analyses of healthcare access and experiences within these populations.

The 2022 HCPS ultimately included interviews with approximately 4,600 patients across 142 health centers. A final sample of 4,414 complete surveys was included in the 2022 public use file. A single composite survey weight was applied to each response to account for the complex sampling design, including unequal probabilities of selection and nonresponse adjustments. These weights enable valid population-level estimates reflective of the entire Health Center Program patient population nationwide.

### Measures

The Health Center Patient Survey (HCPS) collects detailed information across multiple domains relevant to healthcare access, utilization, and outcomes among patients served by HRSA-funded health centers. Core domains include sociodemographic characteristics, health insurance status, usual source of care, health conditions and behaviors, experiences with care, and patient satisfaction. The survey instrument, administered via in-person interviews, emphasizes patient-reported experiences and self-identified health needs.

#### Demographic and Clinic Characteristics

Demographic measures include patient age, sex, race, ethnicity, educational attainment, income, employment, housing status, language spoken at home, and country of birth. The survey also asks about health insurance coverage and self-reported healthcare access barriers. Patients are categorized by the type of health center they most recently visited, which includes CHCs, HCH clinics, and PHPC programs. This information allows stratified analyses by health center designation, particularly important for understanding care delivery to vulnerable populations.

#### Indicators Aligned with Life’s Essential 8

To assess cardiovascular health in alignment with the AHA’s LE8 framework, several self-reported items from the HCPS were utilized. Sleep: Average hours of sleep on a typical night was recorded, allowing classification consistent with LE8 sleep health guidelines (7-9 hours) and characterization of the distribution; Physical Activity: Respondents were asked about the frequency and duration of moderate and vigorous physical activity in a typical week; Smoking: Current tobacco use (cigarettes and other tobacco products) was documented; Nutrition: Dietary concerns were assessed through questions about receiving advice related to diet from a doctor or other health professional; Body Mass Index (BMI): Calculated from self-reported height and weight; Cholesterol: Respondents were asked whether they had ever been told by a health professional that they had high cholesterol; Blood Pressure/Hypertension: Self-reported whether a health care provider had ever informed the respondent of hypertension or high-blood pressure; Blood Sugar/Diabetes: self-reported diagnosis by a clinician, including both type 1 and type 2 diabetes. The survey also asked about A1c testing and results, where available. These measures allow for at least partial capture and construction of LE8-based profiles and support examining cardiovascular health status among diverse populations served by these two types of federally funded health centers.

### Analysis

The data were cleaned to remove missing response data, including survey responses of “don’t know” and “refusal,” which were treated as missing. Demographics were characterized for the entire sample by frequency, percentage, and then the survey-weighted percent. For CHC and HCH, survey-weighted frequencies and percentages were calculated for all LE8-related item responses. The proportions of responses were tested for independence between clinic types and bivariate logistic regression to estimate effect sizes (odds ratios, 95% confidence intervals) between HCH clinic access and all items.The LE8 characteristics were then compared between CHC and HCH using multivariate logistic regression, adjusting for age, sex, race/ethnicity, language preference, marital status, and employment status.

## Results

As seen in Table 1 above, using survey-weighted percentages, the sample was predominantly female (57%), non-Hispanic White (36%), and aged 18-44 years old (36%). Overall, 65% did not speak another language at home, the majority were not veterans (97%), approximately half were not in the labor force (48%), were married or had a domestic partner (44%), and had never been homeless (87%). Most of the sample received services at the reference CHCs (90%). For the cardiovascular factors, 59% of the sample smoked less than 100 cigarettes in a lifetime, 79% had at least one day a week of physical activity, 74% were not told they had a weight problem within the past year by a healthcare provider, and 93% had not been diagnosed with CVD.

**Table 1.** Sample Demographics.

|  | Sample n<br>(n = 4414*) | Sample<br>Percent (%) | Survey-Weighted<br>Percent (%) |
| --- | --- | --- | --- |
| **<u>Sex</u>** |  |  |  |
| Male | 1477 | 33.46 | 42.65 |
| Female | 2937 | 66.54 | 57.35 |
| **<u>Race</u>** |  |  |  |
| Non-Hispanic White | 1055 | 23.9 | 35.94 |
| Hispanic | 1869 | 42.34 | 32.27 |
| Non-Hispanic Black | 886 | 20.07 | 16.41 |
| Other | 604 | 13.68 | 15.38 |
| **<u>Ethnicity</u>** |  |  |  |
| Non-Hispanic | 2477 | 56.23 | 65.39 |
| Hispanic | 1928 | 43.77 | 34.61 |
| **<u>Age</u>** |  |  |  |
| 0-17 years | 502 | 11.37 | 28.81 |
| 18-44 years | 1372 | 31.08 | 35.79 |
| 45-64 years | 1889 | 42.8 | 24.55 |
| 65-74 years | 526 | 11.92 | 8.7 |
| 75+ years | 125 | 2.83 | 2.15 |
| **<u>Language Other than English at Home</u>** |  |  |  |
| No | 2611 | 59.21 | 64.87 |
| Yes | 1799 | 40.79 | 35.13 |
| **<u>Veteran Status</u>** |  |  |  |
| No | 3779 | 96.9 | 97.05 |
| Yes | 121 | 3.1 | 2.95 |
| **<u>Employment Status</u>** |  |  |  |
| Unemployed | 501 | 11.35 | 9.71 |
| Employed | 1560 | 35.34 | 42.57 |
| Not in the labor force | 2353 | 53.31 | 47.72 |
| **<u>Marital Status</u>** |  |  |  |
| Never been married | 1247 | 32.11 | 34.87 |
| Married or have a domestic partner | 1479 | 38.08 | 43.79 |
| Widowed, divorced, or separated | 1158 | 29.81 | 21.34 |
| **<u>Homelessness (ever)</u>** |  |  |  |
| No | 3097 | 84.97 | 86.53 |
| Yes | 548 | 15.03 | 13.47 |
| **<u>Smoking Status (at least 100 cigarettes in a lifetime)</u>** |  |  |  |
| No | 2384 | 59.96 | 58.82 |
| Yes | 1592 | 40.04 | 41.18 |
| **<u>Physical Activity in the Past 7 days</u>** |  |  |  |
| No | 1069 | 24.52 | 21.13 |
| Yes (1 day or more) | 3291 | 75.48 | 78.87 |
| **<u>Average Hours of Sleep</u>** |  |  |  |
| Less than 8 hours | 2436 | 55.45 | 45.35 |
| 8 hours | 1245 | 28.34 | 30.11 |
| Greater than 8 hours | 712 | 16.21 | 24.54 |
| **<u>Told Weight Problem by a Healthcare Professional (within the past year)</u>** |  |  |  |
| No | 3110 | 70.68 | 74.26 |
| Yes | 1290 | 29.32 | 25.74 |
| **<u>Patient Type</u>** |  |  |  |
| Public Housing Primary Care | 429 | 9.72 | 3.3 |
| Migrant Health Center | 473 | 10.72 | 3.82 |
| Health Care for the Homeless | 597 | 13.53 | 2.75 |
| Community Health Center | 2914 | 66.03 | 90.13 |
| **<u>Cardiovascular Disease Status</u>** |  |  |  |
| No | 3983 | 90.36 | 92.5 |
| Yes | 425 | 9.64 | 7.5 |

\*Responses listed as “Don’t Know” and “Refusal” were treated as missing data

As seen in Table 2 above, comparing CHC and HCH clients showed a significant difference in demographic characteristics for average sex, age, language other than English at home, marital status, employment status, and if they had ever been homeless. Those using HCH clinics had 2.02 (95% CI:1.31,3.12) times the odds of being male, 0.096 (95% CI: 0.03, 0.27) of being a child (0-17 years old), and 0.037 (95% CI: 0.005, 0.30) times the odds of being older than 75 years old when compared to CHC clients. Those at HCH clinics had 0.517 (95% CI: 0.32,0.84) times the odds of speaking a language other than English at home, 4.309 (95% CI: 2.62, 7.08) times of never being married, and 4.474 (95% CI: 2.85, 7.84) times the odds of being widowed, divorced, or separated when compared to CHC users. HCH clinic patients had 5.004 (95% CI: 2.86,8.90) times the odds of being unemployed and 12.562 (95% CI: 4.58, 34.42) times the odds of ever being homeless compared to CHC patients.

**Table 2.**
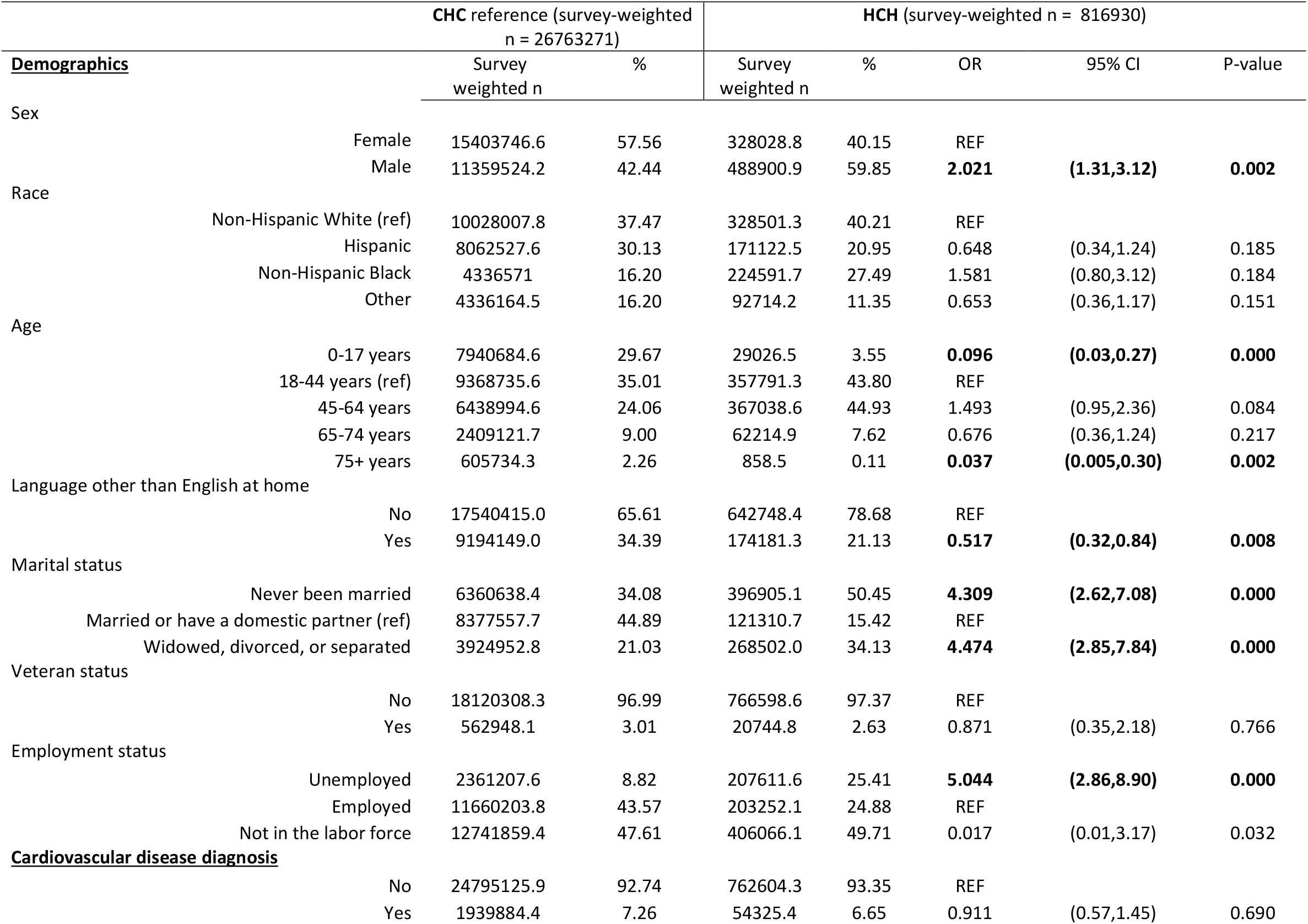
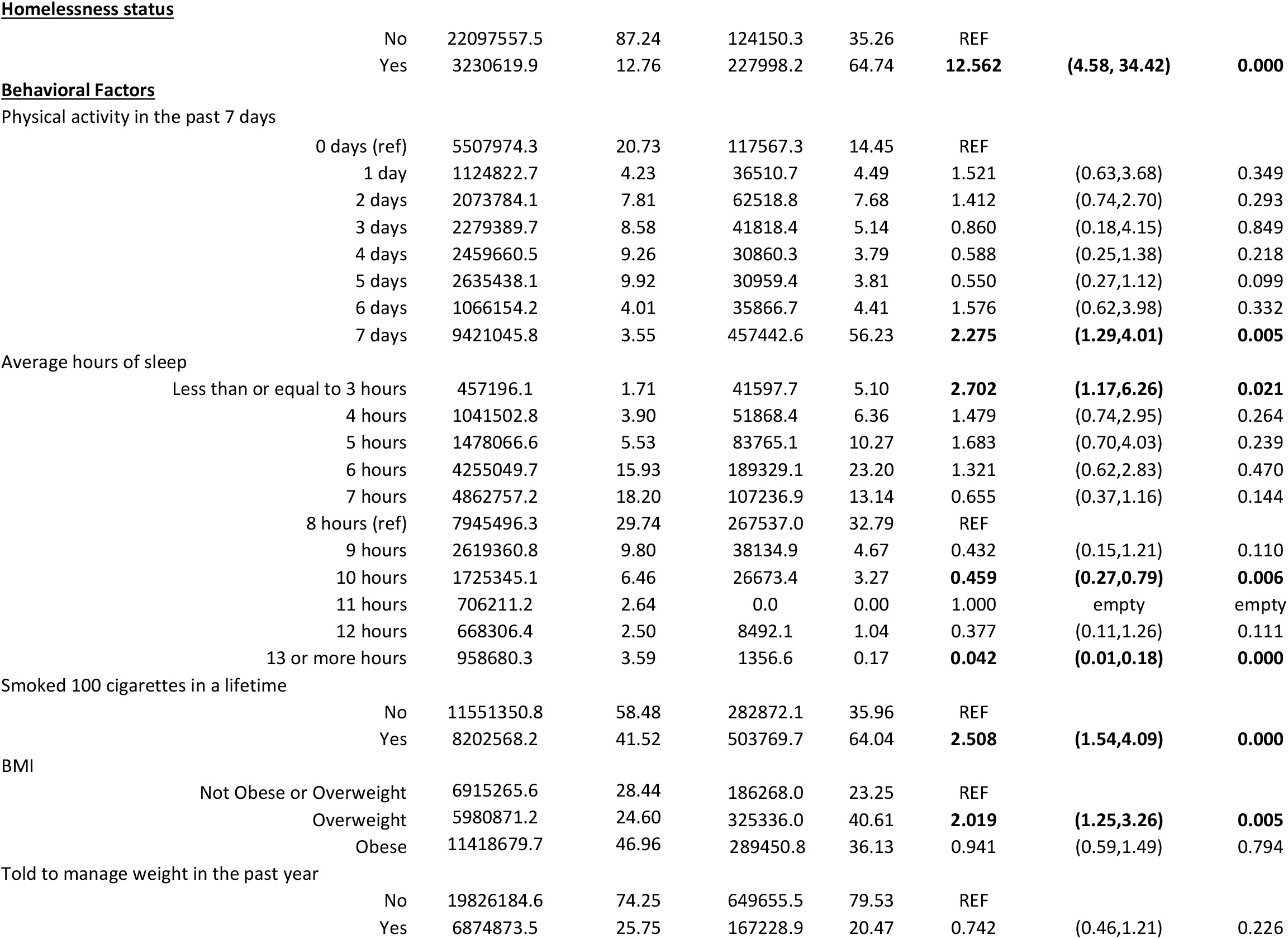
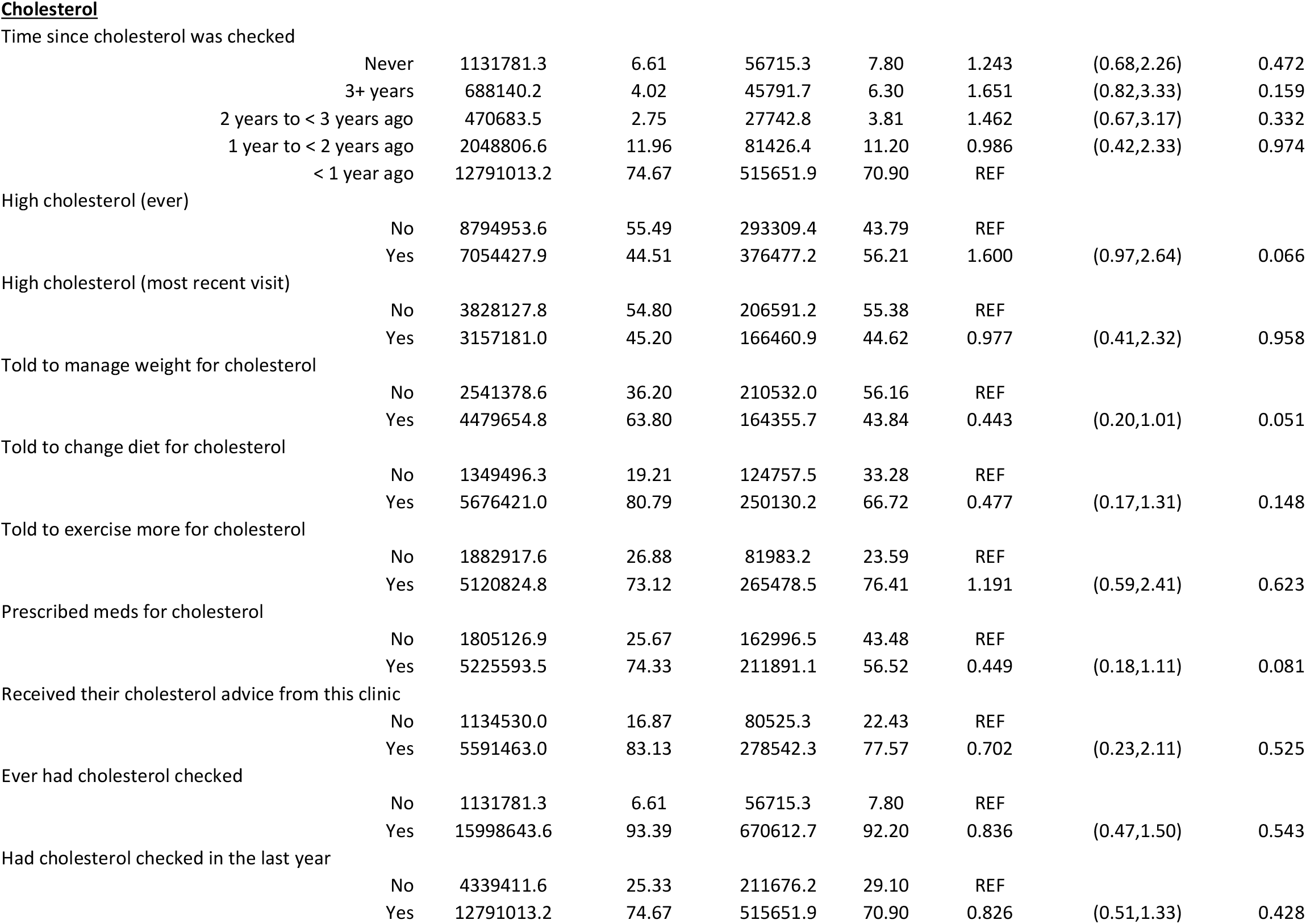
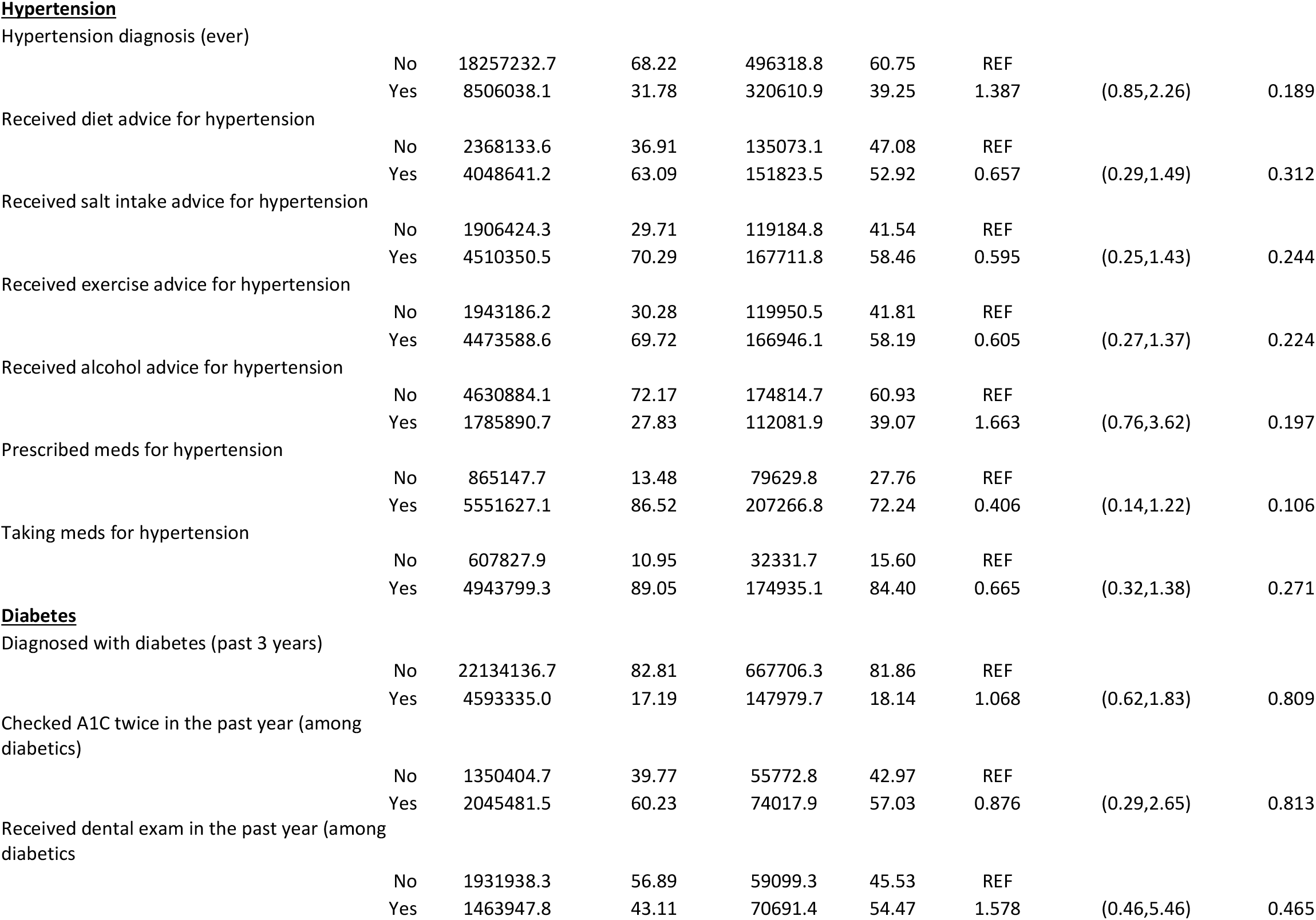
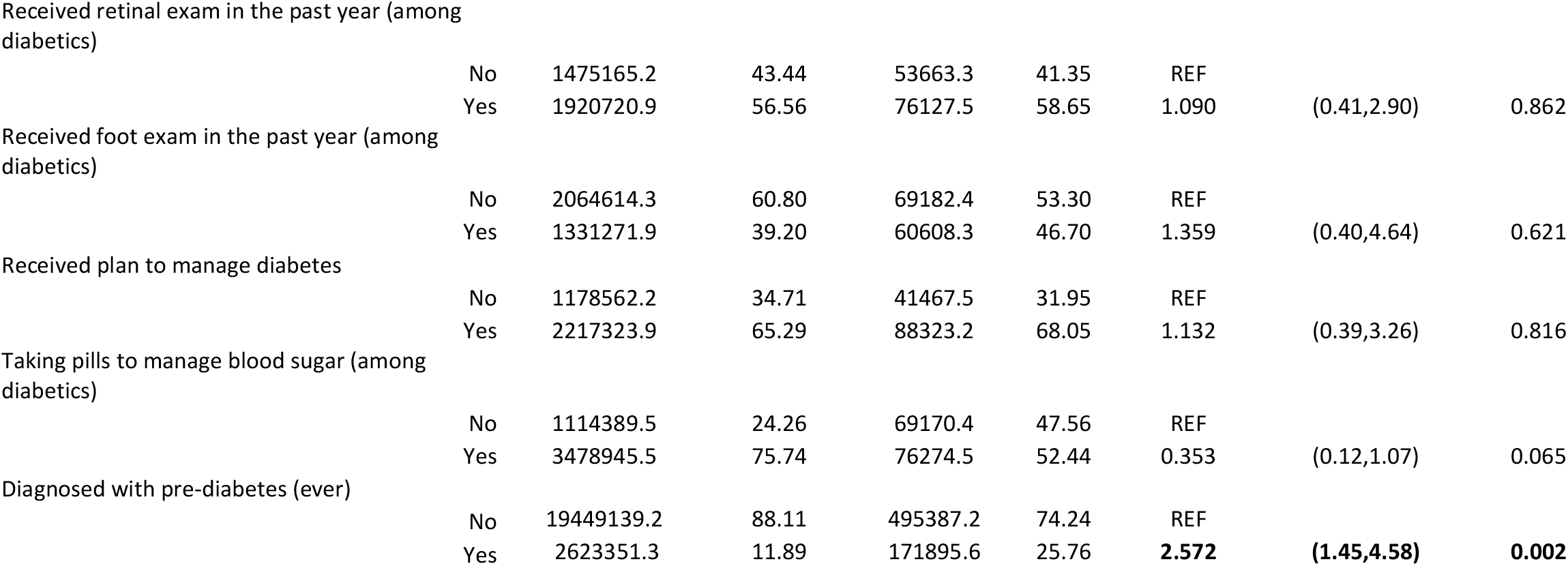
Survey Weighted LE8 between Health Care for the Homeless and Community Health Center.

|  |  | CHC reference (survey-weighted<br>n = 26763271) |  | HCH (survey-weighted n = 816930) |  |  |  |  |
| --- | --- | --- | --- | --- | --- | --- | --- | --- |
| <u>Demographics</u> |  | Survey<br>weighted n | % | Survey<br>weighted n | % | OR | 95% CI | P-value |
| Sex |  |  |  |  |  |  |  |  |
|  | Female | 15403746.6 | 57.56 | 328028.8 | 40.15 | REF |  |  |
|  | Male | 11359524.2 | 42.44 | 488900.9 | 59.85 | **2.021** | **(1.31,3.12)** | **0.002** |
| Race |  |  |  |  |  |  |  |  |
|  | Non-Hispanic White (ref) | 10028007.8 | 37.47 | 328501.3 | 40.21 | REF |  |  |
|  | Hispanic | 8062527.6 | 30.13 | 171122.5 | 20.95 | 0.648 | (0.34,1.24) | 0.185 |
|  | Non-Hispanic Black | 4336571 | 16.20 | 224591.7 | 27.49 | 1.581 | (0.80,3.12) | 0.184 |
|  | Other | 4336164.5 | 16.20 | 92714.2 | 11.35 | 0.653 | (0.36,1.17) | 0.151 |
| Age |  |  |  |  |  |  |  |  |
|  | 0-17 years | 7940684.6 | 29.67 | 29026.5 | 3.55 | **0.096** | **(0.03,0.27)** | **0.000** |
|  | 18-44 years (ref) | 9368735.6 | 35.01 | 357791.3 | 43.80 | REF |  |  |
|  | 45-64 years | 6438994.6 | 24.06 | 367038.6 | 44.93 | 1.493 | (0.95,2.36) | 0.084 |
|  | 65-74 years | 2409121.7 | 9.00 | 62214.9 | 7.62 | 0.676 | (0.36,1.24) | 0.217 |
|  | 75+ years | 605734.3 | 2.26 | 858.5 | 0.11 | **0.037** | **(0.005,0.30)** | **0.002** |
| Language other than English at home |  |  |  |  |  |  |  |  |
|  | No | 17540415.0 | 65.61 | 642748.4 | 78.68 | REF |  |  |
|  | Yes | 9194149.0 | 34.39 | 174181.3 | 21.13 | **0.517** | **(0.32,0.84)** | **0.008** |
| Marital status |  |  |  |  |  |  |  |  |
|  | Never been married | 6360638.4 | 34.08 | 396905.1 | 50.45 | **4.309** | **(2.62,7.08)** | **0.000** |
|  | Married or have a domestic partner (ref) | 8377557.7 | 44.89 | 121310.7 | 15.42 | REF |  |  |
|  | Widowed, divorced, or separated | 3924952.8 | 21.03 | 268502.0 | 34.13 | **4.474** | **(2.85,7.84)** | **0.000** |
| Veteran status |  |  |  |  |  |  |  |  |
|  | No | 18120308.3 | 96.99 | 766598.6 | 97.37 | REF |  |  |
|  | Yes | 562948.1 | 3.01 | 20744.8 | 2.63 | 0.871 | (0.35,2.18) | 0.766 |
| Employment status |  |  |  |  |  |  |  |  |
|  | Unemployed | 2361207.6 | 8.82 | 207611.6 | 25.41 | **5.044** | **(2.86,8.90)** | **0.000** |
|  | Employed | 11660203.8 | 43.57 | 203252.1 | 24.88 | REF |  |  |
|  | Not in the labor force | 12741859.4 | 47.61 | 406066.1 | 49.71 | 0.017 | (0.01,3.17) | 0.032 |
| <u>Cardiovascular disease diagnosis</u> |  |  |  |  |  |  |  |  |
|  | No | 24795125.9 | 92.74 | 762604.3 | 93.35 | REF |  |  |
|  | Yes | 1939884.4 | 7.26 | 54325.4 | 6.65 | 0.911 | (0.57,1.45) | 0.690 |

Homelessness status
|  |  |  |  |  |  |  |  |
| --- | --- | --- | --- | --- | --- | --- | --- |
| No | 22097557.5 | 87.24 | 124150.3 | 35.26 | REF |  |  |
| Yes | 3230619.9 | 12.76 | 227998.2 | 64.74 | **12.562** | **(4.58, 34.42)** | **0.000** |

Physical activity in the past 7 days
|  |  |  |  |  |  |  |  |
| --- | --- | --- | --- | --- | --- | --- | --- |
| 0 days (ref) | 5507974.3 | 20.73 | 117567.3 | 14.45 | REF |  |  |
| 1 day | 1124822.7 | 4.23 | 36510.7 | 4.49 | 1.521 | (0.63,3.68) | 0.349 |
| 2 days | 2073784.1 | 7.81 | 62518.8 | 7.68 | 1.412 | (0.74,2.70) | 0.293 |
| 3 days | 2279389.7 | 8.58 | 41818.4 | 5.14 | 0.860 | (0.18,4.15) | 0.849 |
| 4 days | 2459660.5 | 9.26 | 30860.3 | 3.79 | 0.588 | (0.25,1.38) | 0.218 |
| 5 days | 2635438.1 | 9.92 | 30959.4 | 3.81 | 0.550 | (0.27,1.12) | 0.099 |
| 6 days | 1066154.2 | 4.01 | 35866.7 | 4.41 | 1.576 | (0.62,3.98) | 0.332 |
| 7 days | 9421045.8 | 3.55 | 457442.6 | 56.23 | **2.275** | **(1.29,4.01)** | **0.005** |

Average hours of sleep
|  |  |  |  |  |  |  |  |
| --- | --- | --- | --- | --- | --- | --- | --- |
| Less than or equal to 3 hours | 457196.1 | 1.71 | 41597.7 | 5.10 | **2.702** | **(1.17,6.26)** | **0.021** |
| 4 hours | 1041502.8 | 3.90 | 51868.4 | 6.36 | 1.479 | (0.74,2.95) | 0.264 |
| 5 hours | 1478066.6 | 5.53 | 83765.1 | 10.27 | 1.683 | (0.70,4.03) | 0.239 |
| 6 hours | 4255049.7 | 15.93 | 189329.1 | 23.20 | 1.321 | (0.62,2.83) | 0.470 |
| 7 hours | 4862757.2 | 18.20 | 107236.9 | 13.14 | 0.655 | (0.37,1.16) | 0.144 |
| 8 hours (ref) | 7945496.3 | 29.74 | 267537.0 | 32.79 | REF |  |  |
| 9 hours | 2619360.8 | 9.80 | 38134.9 | 4.67 | 0.432 | (0.15,1.21) | 0.110 |
| 10 hours | 1725345.1 | 6.46 | 26673.4 | 3.27 | **0.459** | **(0.27,0.79)** | **0.006** |
| 11 hours | 706211.2 | 2.64 | 0.0 | 0.00 | 1.000 | empty | empty |
| 12 hours | 668306.4 | 2.50 | 8492.1 | 1.04 | 0.377 | (0.11,1.26) | 0.111 |
| 13 or more hours | 958680.3 | 3.59 | 1356.6 | 0.17 | **0.042** | **(0.01,0.18)** | **0.000** |

Smoked 100 cigarettes in a lifetime
|  |  |  |  |  |  |  |  |
| --- | --- | --- | --- | --- | --- | --- | --- |
| No | 11551350.8 | 58.48 | 282872.1 | 35.96 | REF |  |  |
| Yes | 8202568.2 | 41.52 | 503769.7 | 64.04 | **2.508** | **(1.54,4.09)** | **0.000** |

BMI
|  |  |  |  |  |  |  |  |
| --- | --- | --- | --- | --- | --- | --- | --- |
| Not Obese or Overweight | 6915265.6 | 28.44 | 186268.0 | 23.25 | REF |  |  |
| Overweight | 5980871.2 | 24.60 | 325336.0 | 40.61 | **2.019** | **(1.25,3.26)** | **0.005** |
| Obese | 11418679.7 | 46.96 | 289450.8 | 36.13 | 0.941 | (0.59,1.49) | 0.794 |

Told to manage weight in the past year
|  |  |  |  |  |  |  |  |
| --- | --- | --- | --- | --- | --- | --- | --- |
| No | 19826184.6 | 74.25 | 649655.5 | 79.53 | REF |  |  |
| Yes | 6874873.5 | 25.75 | 167228.9 | 20.47 | 0.742 | (0.46,1.21) | 0.226 |

Time since cholesterol was checked
|  |  |  |  |  |  |  |  |
| --- | --- | --- | --- | --- | --- | --- | --- |
| Never | 1131781.3 | 6.61 | 56715.3 | 7.80 | 1.243 | (0.68,2.26) | 0.472 |
| 3+ years | 688140.2 | 4.02 | 45791.7 | 6.30 | 1.651 | (0.82,3.33) | 0.159 |
| 2 years to < 3 years ago | 470683.5 | 2.75 | 27742.8 | 3.81 | 1.462 | (0.67,3.17) | 0.332 |
| 1 year to < 2 years ago | 2048806.6 | 11.96 | 81426.4 | 11.20 | 0.986 | (0.42,2.33) | 0.974 |
| < 1 year ago | 12791013.2 | 74.67 | 515651.9 | 70.90 | REF |  |  |

High cholesterol (ever)
|  |  |  |  |  |  |  |  |
| --- | --- | --- | --- | --- | --- | --- | --- |
| No | 8794953.6 | 55.49 | 293309.4 | 43.79 | REF |  |  |
| Yes | 7054427.9 | 44.51 | 376477.2 | 56.21 | 1.600 | (0.97,2.64) | 0.066 |

High cholesterol (most recent visit)
|  |  |  |  |  |  |  |  |
| --- | --- | --- | --- | --- | --- | --- | --- |
| No | 3828127.8 | 54.80 | 206591.2 | 55.38 | REF |  |  |
| Yes | 3157181.0 | 45.20 | 166460.9 | 44.62 | 0.977 | (0.41,2.32) | 0.958 |

Told to manage weight for cholesterol
|  |  |  |  |  |  |  |  |
| --- | --- | --- | --- | --- | --- | --- | --- |
| No | 2541378.6 | 36.20 | 210532.0 | 56.16 | REF |  |  |
| Yes | 4479654.8 | 63.80 | 164355.7 | 43.84 | 0.443 | (0.20,1.01) | 0.051 |

Told to change diet for cholesterol
|  |  |  |  |  |  |  |  |
| --- | --- | --- | --- | --- | --- | --- | --- |
| No | 1349496.3 | 19.21 | 124757.5 | 33.28 | REF |  |  |
| Yes | 5676421.0 | 80.79 | 250130.2 | 66.72 | 0.477 | (0.17,1.31) | 0.148 |

Told to exercise more for cholesterol
|  |  |  |  |  |  |  |  |
| --- | --- | --- | --- | --- | --- | --- | --- |
| No | 1882917.6 | 26.88 | 81983.2 | 23.59 | REF |  |  |
| Yes | 5120824.8 | 73.12 | 265478.5 | 76.41 | 1.191 | (0.59,2.41) | 0.623 |

Prescribed meds for cholesterol
|  |  |  |  |  |  |  |  |
| --- | --- | --- | --- | --- | --- | --- | --- |
| No | 1805126.9 | 25.67 | 162996.5 | 43.48 | REF |  |  |
| Yes | 5225593.5 | 74.33 | 211891.1 | 56.52 | 0.449 | (0.18,1.11) | 0.081 |

Received their cholesterol advice from this clinic
|  |  |  |  |  |  |  |  |
| --- | --- | --- | --- | --- | --- | --- | --- |
| No | 1134530.0 | 16.87 | 80525.3 | 22.43 | REF |  |  |
| Yes | 5591463.0 | 83.13 | 278542.3 | 77.57 | 0.702 | (0.23,2.11) | 0.525 |

Ever had cholesterol checked
|  |  |  |  |  |  |  |  |
| --- | --- | --- | --- | --- | --- | --- | --- |
| No | 1131781.3 | 6.61 | 56715.3 | 7.80 | REF |  |  |
| Yes | 15998643.6 | 93.39 | 670612.7 | 92.20 | 0.836 | (0.47,1.50) | 0.543 |

Had cholesterol checked in the last year
|  |  |  |  |  |  |  |  |
| --- | --- | --- | --- | --- | --- | --- | --- |
| No | 4339411.6 | 25.33 | 211676.2 | 29.10 | REF |  |  |
| Yes | 12791013.2 | 74.67 | 515651.9 | 70.90 | 0.826 | (0.51,1.33) | 0.428 |

|  |  |  |  |  |  |  |  |
| --- | --- | --- | --- | --- | --- | --- | --- |
| No | 18257232.7 | 68.22 | 496318.8 | 60.75 | REF |  |  |
| Yes | 8506038.1 | 31.78 | 320610.9 | 39.25 | 1.387 | (0.85,2.26) | 0.189 |

Received diet advice for hypertension
|  |  |  |  |  |  |  |  |
| --- | --- | --- | --- | --- | --- | --- | --- |
| No | 2368133.6 | 36.91 | 135073.1 | 47.08 | REF |  |  |
| Yes | 4048641.2 | 63.09 | 151823.5 | 52.92 | 0.657 | (0.29,1.49) | 0.312 |

Received salt intake advice for hypertension
|  |  |  |  |  |  |  |  |
| --- | --- | --- | --- | --- | --- | --- | --- |
| No | 1906424.3 | 29.71 | 119184.8 | 41.54 | REF |  |  |
| Yes | 4510350.5 | 70.29 | 167711.8 | 58.46 | 0.595 | (0.25,1.43) | 0.244 |

Received exercise advice for hypertension
|  |  |  |  |  |  |  |  |
| --- | --- | --- | --- | --- | --- | --- | --- |
| No | 1943186.2 | 30.28 | 119950.5 | 41.81 | REF |  |  |
| Yes | 4473588.6 | 69.72 | 166946.1 | 58.19 | 0.605 | (0.27,1.37) | 0.224 |

Received alcohol advice for hypertension
|  |  |  |  |  |  |  |  |
| --- | --- | --- | --- | --- | --- | --- | --- |
| No | 4630884.1 | 72.17 | 174814.7 | 60.93 | REF |  |  |
| Yes | 1785890.7 | 27.83 | 112081.9 | 39.07 | 1.663 | (0.76,3.62) | 0.197 |

Prescribed meds for hypertension
|  |  |  |  |  |  |  |  |
| --- | --- | --- | --- | --- | --- | --- | --- |
| No | 865147.7 | 13.48 | 79629.8 | 27.76 | REF |  |  |
| Yes | 5551627.1 | 86.52 | 207266.8 | 72.24 | 0.406 | (0.14,1.22) | 0.106 |

Taking meds for hypertension
|  |  |  |  |  |  |  |  |
| --- | --- | --- | --- | --- | --- | --- | --- |
| No | 607827.9 | 10.95 | 32331.7 | 15.60 | REF |  |  |
| Yes | 4943799.3 | 89.05 | 174935.1 | 84.40 | 0.665 | (0.32,1.38) | 0.271 |

|  |  |  |  |  |  |  |  |
| --- | --- | --- | --- | --- | --- | --- | --- |
| No | 22134136.7 | 82.81 | 667706.3 | 81.86 | REF |  |  |
| Yes | 4593335.0 | 17.19 | 147979.7 | 18.14 | 1.068 | (0.62,1.83) | 0.809 |

Checked A1C twice in the past year (among diabetics)
|  |  |  |  |  |  |  |  |
| --- | --- | --- | --- | --- | --- | --- | --- |
| No | 1350404.7 | 39.77 | 55772.8 | 42.97 | REF |  |  |
| Yes | 2045481.5 | 60.23 | 74017.9 | 57.03 | 0.876 | (0.29,2.65) | 0.813 |

Received dental exam in the past year (among diabetics)
|  |  |  |  |  |  |  |  |
| --- | --- | --- | --- | --- | --- | --- | --- |
| No | 1931938.3 | 56.89 | 59099.3 | 45.53 | REF |  |  |
| Yes | 1463947.8 | 43.11 | 70691.4 | 54.47 | 1.578 | (0.46,5.46) | 0.465 |
| Received retinal exam in the past year (among diabetics) | No | 1475165.2 | 43.44 | 53663.3 | 41.35 | REF |  |  |
|  | Yes | 1920720.9 | 56.56 | 76127.5 | 58.65 | 1.090 | (0.41,2.90) | 0.862 |
| Received foot exam in the past year (among diabetics) | No | 2064614.3 | 60.80 | 69182.4 | 53.30 | REF |  |  |
|  | Yes | 1331271.9 | 39.20 | 60608.3 | 46.70 | 1.359 | (0.40,4.64) | 0.621 |
| Received plan to manage diabetes | No | 1178562.2 | 34.71 | 41467.5 | 31.95 | REF |  |  |
|  | Yes | 2217323.9 | 65.29 | 88323.2 | 68.05 | 1.132 | (0.39,3.26) | 0.816 |
| Taking pills to manage blood sugar (among diabetics) | No | 1114389.5 | 24.26 | 69170.4 | 47.56 | REF |  |  |
|  | Yes | 3478945.5 | 75.74 | 76274.5 | 52.44 | 0.353 | (0.12,1.07) | 0.065 |
| Diagnosed with pre-diabetes (ever) | No | 19449139.2 | 88.11 | 495387.2 | 74.24 | REF |  |  |
|  | Yes | 2623351.3 | 11.89 | 171895.6 | 25.76 | **2.572** | **(1.45,4.58)** | **0.002** |

The LE8 factors between CHC and HCH significantly differed in levels of physical activity, average hours of sleep, smoking status, body mass index (BMI), and pre-diabetes diagnosis. HCH clients had 2.702 (95% CI: 1.17, 6.26) times the odds of being physically active on all seven days of the week on average compared to CHC clients. In terms of average hours of sleep a night, HCH patients had 2.702 (95% CI: 1.17,6.26) times the odds of getting less than or equal to three hours,0.459 (95% CI: 0.27,0.79) times the odds of getting ten hours, and 0.042 (0.01,0.18) times the odds of getting 13 or more hours when compared to CHC. Participants using HCH had 2.508 (95% CI: 1.54,4.09) times the odds of having smoked greater than 100 cigarettes in their lifetime compared to CHC participants. For BMI, HCH patients had 2.019 (95% CI: 1.25,3.26) times the odds of being overweight (BMI of 25-29.9) and additionally had 2.572 (95% CI: 1.45,4.58) times the odds of ever being diagnosed with pre-diabetes when compared to CHC patients. All other measures for the LE8 were found not to be statistically significantly different between HCH and CHC.

Table 3 shows the differences between HCH and CHC remain for the average level of physical activity, average hours of sleep, smoking history, and pre-diabetes diagnosis after adjusting for age, sex, race/ethnicity, language preference, marital status, and employment status. HCH clients had an average of 2.686 (95% CI: 1.43,5.04) times the odds of being physically active every day of the week compared to CHC clients after adjustment. The multivariable models also showed that those using HCH clinics had 0.487 (95% CI: 0.27,0.88) times the odds of getting an average of 7 hours of sleep a night and 0.075 (95% CI: 0.01,0.42) times the odds of getting greater than 13 hours a of sleep a night on average compared to CHC after adjustment. Those utilizing HCH clinics had 2.07 (95% CI: 1.24,3.46) times the odds of smoking at least 100 cigarettes in their lifetime and 2.29 (95% CI: 1.19,4.40) times the odds of being diagnosed as pre-diabetic when compared to those using CHC clinics, after adjusting for demographics and socioeconomic measures.

**Table 3.** Survey-Weighted Logistic Regression Model*.

|  |  | HCH (CHC as reference) |  |  |
| --- | --- | --- | --- | --- |
|  |  | OR | 95% CI | p-value |
| **<u>Behavioral Factors</u>** |  |  |  |  |
| Physical activity in the past 7 days |  |  |  |  |
|  | 0 days | REF | REF | REF |
|  | 1 day | 1.076 | (0.41,0.80) | 0.879 |
|  | 2 days | 1.594 | (0.84,3.02) | 0.151 |
|  | 3 days | 0.855 | (0.18,4.04) | 0.842 |
|  | 4 days | 0.615 | (0.24,1.55) | 0.300 |
|  | 5 days | 0.546 | (0.24,1.25) | 0.150 |
|  | 6 days | 1.868 | (0.65,5.35) | 0.242 |
|  | 7 days | **2.686** | **(1.43,5.04)** | **0.002** |
| Average hours of sleep |  |  |  |  |
|  | ≤3 hours | 1.292 | (0.38,4.36) | 0.676 |
|  | 4 hours | 0.871 | (0.42,1.82) | 0.711 |
|  | 5 hours | 1.078 | (0.41,2.83) | 0.878 |
|  | 6 hours | 0.703 | (0.33,1.49) | 0.353 |
|  | 7 hours | **0.487** | **(0.27,0.88)** | **0.017** |
|  | 8 hours | REF | REF | REF |
|  | 9 hours | 0.532 | (0.22,1.28) | 0.158 |
|  | 10 hours | 0.737 | (0.28,1.94) | 0.531 |
|  | 11 hours | 1.000 | empty |  |
|  | 12 hours | 0.362 | (0.07,1.86) | 0.220 |
|  | 13 + hours | **0.075** | **(0.01,0.42)** | **0.004** |
| Smoked 100 cigarettes in a lifetime |  | **2.07** | **(1.24,3.46)** | **0.006** |
| BMI |  |  |  |  |
|  | Not Obese or Overweight | REF | REF | REF |
|  | Overweight | 1.491 | (0.88,2.53) | 0.138 |
|  | Obese | 0.836 | (0.53,1.33) | 0.445 |
| **<u>Cholesterol</u>** |  |  |  |  |
|  | High cholesterol (ever) | 1.710 | (0.99,2.96) | 0.055 |
|  | High cholesterol (most recent visit) | 1.596 | (0.74,3.42) | 0.227 |
| **<u>Hypertension</u>** |  |  |  |  |
|  | Hypertension diagnosis (ever) | 0.567 | (0.30,1.08) | 0.083 |
| **<u>Diabetes</u>** |  |  |  |  |
|  | Diagnosed with diabetes (past 3 years) | 0.631 | (0.38,1.04) | 0.073 |
|  | Diagnosed with pre-diabetes ever | **2.291** | **(1.19, 4.40)** | **0.014** |

\*Model adjusted for age, sex, race/ethnicity, language preference, marital status, and employment status

## Discussion

This analysis of the 2022 Health Center Patient Survey (HCPS) provides important insights into the cardiovascular health profiles of patients served by Health Care for the Homeless (HCH) clinics compared to those seen at Community Health Centers (CHCs), using the AHA’s LE8 framework. While both clinic types operate within HRSA’s Health Center Program and serve medically underserved populations, our findings underscore how the social and structural vulnerabilities of HCH patients result in distinctive risk patterns for cardiovascular disease (CVD).

Notably, patients seen in HCH settings differ significantly in key sociodemographic characteristics. They were more likely to be male, unemployed, unmarried, and to have a personal history of homelessness. These factors reflect the intended target population of HCH clinics and highlight the complexity of care delivery in these settings. The housing insecurity and instability experienced by many HCH patients can influence their exposure to health risks and their ability to maintain long-term engagement with preventive care.

Across LE8 measures, HCH patients exhibited a mixed cardiovascular risk profile. They were more likely than CHC patients to report daily physical activity. Yet, they also had significantly higher odds of inadequate sleep duration, a history of smoking, and a pre-diabetes diagnosis. These findings persisted even after adjusting for demographic and socioeconomic factors, suggesting that the elevated risk in some domains is not solely attributable to baseline differences in population characteristics. Interestingly, HCH patients did not show significantly higher odds of diagnosed hypertension, high cholesterol, or diagnosed diabetes, which may reflect limitations in screening, underdiagnosis, or distinct disease trajectories in this population.

These findings should not be interpreted as reflecting lower quality of care at HCH clinics. Rather, they point to the distinct epidemiological context in which HCH providers operate. Traditional CVD risk assessment tools and prevention models, which rely on stable patterns of healthcare access and biomedical risk markers, may not fully capture the realities of cardiovascular disease progression among people experiencing homelessness. For example, interrupted sleep, high rates of tobacco use, and fluctuating dietary quality may contribute to metabolic and vascular stress in ways that are not immediately evident in standard clinical screenings.

From an implementation perspective, our results support calls for tailored approaches to cardiovascular screening and prevention in HCH settings. Risk stratification algorithms may need recalibration to reflect this population’s complex interplay of behavioral, environmental, and social exposures. Additionally, routine clinical metrics—such as BMI or A1c—may have different prognostic values in chronic stress, exposure to the elements, and fragmented care access. The higher odds of pre-diabetes among HCH patients may represent an early signal of metabolic vulnerability that, if unaddressed, could lead to more severe disease progression over time.

This study contributes to a growing body of literature arguing that housing status is a critical determinant of cardiovascular health. It also reinforces the need for integrated, context-sensitive models of care delivery that account for the cumulative impact of homelessness and structural inequity. HCH clinics are uniquely positioned to provide such care, and future research should explore how to strengthen and adapt evidence-based cardiovascular interventions for this population.

### Limitations

These findings should be interpreted with several limitations in mind. First, the HCPS relies on self-reported data, which may be subject to recall or social desirability bias—especially for sensitive information such as tobacco use, weight, or disease diagnoses. Second, the cross-sectional nature of the survey limits causal inference about the relationship between homelessness, clinic type, and cardiovascular health. Third, while survey weights help generalize to the broader Health Center Program population, the relatively small proportion of HCH respondents may limit statistical power for some subgroup analyses. Finally, certain LE8 components—such as diet quality and cholesterol levels—were assessed using proxy or self-report measures rather than clinical biomarkers, which may limit comparability to any studies using clinical data.

## Conclusions

This study demonstrates that patients accessing Health Care for the Homeless clinics have distinct cardiovascular risk factor profiles compared to those seen at general Community Health Centers, even after accounting for differences in sociodemographic and socioeconomic characteristics. The findings call for a nuanced approach to cardiovascular disease prevention and management among individuals experiencing homelessness, one that recognizes the limitations of standard screening tools and the need for care models tailored to the realities of housing instability and the exposures associated with the experience of homelessness. As the health care system seeks to advance health equity, aligning cardiovascular care strategies with the lived experiences of HCH patients will be essential.

## Acknowledgments

We want to acknowledge the work of the HRSA Bureau of Primary Health Care who conducted and provided the Health Center Patient Survey data for public use. We also thank the respondents to the survey for their contributions to the evidence relied upon.

## Data Availability

The data that support the findings of this study are publicly available from the Health Resources and Services Administration 2022 Health Center Patient Survey. These data can be accessed via the HRSA data portal (https://data.hrsa.gov/topics/health-centers/hcps/dashboard-2022). The HCPS public use file is de-identified and made available for statistical analysis and reporting purposes.

## Author contributions

CRediT: Conceptualization (BK,DB); Methodology (BK,SA); Validation (OJ,SA); Formal analysis (BK,OJ,SA); Investigation (BK,BB,DB); Resources (DB); Data curation (BK,SA); Writing – Original Draft (BK,EC); Writing – Review & Editing (BK,BB,OJ,EC,SA,DB); Supervision (BB,DB); Project administration (BK); Funding acquisition (BK);

